# Innate immune deficiencies in patients with COVID-19

**DOI:** 10.1101/2021.03.29.21254560

**Authors:** Marine Peyneau, Vanessa Granger, Paul-Henri Wicky, Dounia Khelifi-Touhami, Jean-François Timsit, François-Xavier Lescure, Yazdan Yazdanpanah, Alexy Tran-Dihn, Philippe Montravers, Renato C. Monteiro, Sylvie Chollet-Martin, Margarita Hurtado-Nedelec, Luc de Chaisemartin

## Abstract

COVID-19 can cause acute respiratory distress syndrome (ARDS), leading to death in a significant number of individuals. Evidence of a strong role of the innate immune system is accumulating, but the precise cells and mechanism involved remain unclear. In this study, we investigated the links between circulating innate phagocyte phenotype and functions and severity in COVID-19 patients. Eighty-four consecutive patients were included, 44 of which were in intensive care units (ICU). We performed an in-depth phenotyping of neutrophil and monocyte subpopulations and measured soluble activation markers in plasma. Additionally, myeloid cell functions (phagocytosis, oxidative burst, and NETosis) were evaluated on fresh cells from patients. Resulting parameters were linked to disease severity and prognosis. Both ICU and non-ICU patients had circulating neutrophils and monocytes with an activated phenotype, as well as elevated concentrations of soluble activation markers (calprotectin, myeloperoxidase, neutrophil extracellular traps, MMP9, sCD14) in their plasma. ICU patients were characterized by increased CD10^low^ CD13^low^ immature neutrophils, LOX-1^+^ and CCR5^+^ immunosuppressive neutrophils, and HLA-DR^low^ CD14^low^ downregulated monocytes. Markers of immature and immunosuppressive neutrophils were strongly associated with severity and poor outcome. Moreover, neutrophils and monocytes of ICU patients had impaired antimicrobial functions, which correlated with organ dysfunction, severe infections, and mortality. Our study reveals a marked dysregulation of innate immunity in COVID-19 patients, which was correlated with severity and prognosis. Together, our results strongly argue in favor of a pivotal role of innate immunity in COVID-19 severe infections and pleads for targeted therapeutic options.

**One Sentence Summary:** Our study reveals a marked dysregulation of innate immunity in COVID-19 patients, which correlates with severity and prognosis.

## INTRODUCTION

COVID-19 can cause acute respiratory distress syndrome (ARDS), leading to death in a significant number of individuals *(1)*. While the precise mechanisms leading to such a severe outcome are not clear yet, evidence of a strong involvement of the immune system is accumulating *(2, 3)*. High concentrations of proinflammatory cytokines in patients have led to the hypothesis that a “cytokine storm” was one of the main driver of severe COVID-19 *(4)*. While the precise role of cytokines in pathophysiology is debated *(5)*, their circulating concentrations have been associated with severity *(6–8)*, and off-label therapeutic blockade of IL-6 and IL-1 have been attempted with some success *(9–11)*. Furthermore, elevated neutrophil-to-lymphocyte ratio has been linked to severity *(12, 13)*, and neutrophil infiltration has been described in the lungs and broncho-alveolar lavage fluid (BALF) of infected individuals *(14, 15)*. Complexes of DNA and cytoplasmic proteins secreted by activated neutrophils called Neutrophils Extracellular Traps (NETs) have been detected in patients sera *(16)*, and are suspected of participating in lung injury and thrombosis associated with severe COVID-19 *(17– 19)*. Taking into account these findings and previous knowledge on the role of neutrophils in ARDS *(20)*, a major involvement of innate immunity in COVID-19 pathophysiology seems likely *(21, 22)*.

Innate immunity is involved in anti-viral responses. Therefore, in addition to their contribution to hyperinflammation, a dysfunction of innate immunity could also contribute to loss of control of viral replication and thus to COVID-19 severity. Recent data from two French consortia show an impaired type I interferon response in COVID-19 patients associated with persistent viral load *(23–25)*. Moreover, our recent clinical experience and the recent literature report a high incidence (50%) of severe bacterial and fungal infections in non-survivor COVID-19 patients, potentially suggesting an immunosuppression. These infections, in particular invasive aspergillosis, are reminiscent of primary innate immune deficiencies like chronic granulomatous disease *(26)*. Altogether, these findings strongly suggest a role for neutrophils and monocytes in hyperinflammatory responses in COVID-19 pathogenesis, but also in the late immunodeficiency observed in some patients. As highlighted by several authors, there is an urgent need to better document the immune response to be able to both enhance its efficacy and prevent its deleterious effects*(21)*.

In this paper, we investigated the phenotype and functions of circulating phagocytes in hospitalized COVID-19 patients of various severity. Circulating neutrophils and monocytes presented markers of activated cells but also of immature and immunomodulatory subpopulations. At the functional level, phagocytosis, oxidative burst and NETosis were impaired in severe patients. Immature neutrophil markers and functional impairment were linked to organ dysfunction, severe secondary infections, and mortality. These findings indicate that SARS-CoV2 infection may induce an overactivation of the innate system resulting in apparition of abnormal subpopulations and exhaustion of cellular effectors. Such dysregulation of innate responses should be integrated into patient’ s care to help optimizing therapeutic interventions, in particular in severe patients.

## RESULTS

### Neutrophils and monocytes are activated in COVID-19 patients

To assess the activation status of blood phagocytes in COVID-19 patients, we first measured the expression of classical activation markers on both neutrophils and monocytes, as well as soluble activation markers in plasma (Fig.1). Expression of adhesion molecule CD66b was significantly elevated on neutrophils of both non-ICU and ICU patients as compared with healthy controls (Fig.1A), meaning that the cells were activated and underwent degranulation. This was confirmed by the upregulation of CD11b integrin on both patient populations, even if the difference reached statistical significance for non-ICU patients only (Fig.1B). In contrast, L-selectin (CD62L) shedding and FcγRIII (CD16) downregulation were only observed on neutrophils of ICU patients (Fig.1C-D). We confirmed neutrophil activation by measuring cytosolic and granule-derived mediators in the plasma (Fig.1E-G, Fig.S1). Concentrations of MPO and elastase (primary granules), lipocalin-2 (secondary granules), and calprotectin (cytosol) were significantly elevated in both patient groups, while MMP-9 (tertiary granules) was significantly elevated in ICU patients. Additionally, the levels of circulating NETs were strongly increased in both patient groups (Fig.1H).

**Figure 1:**
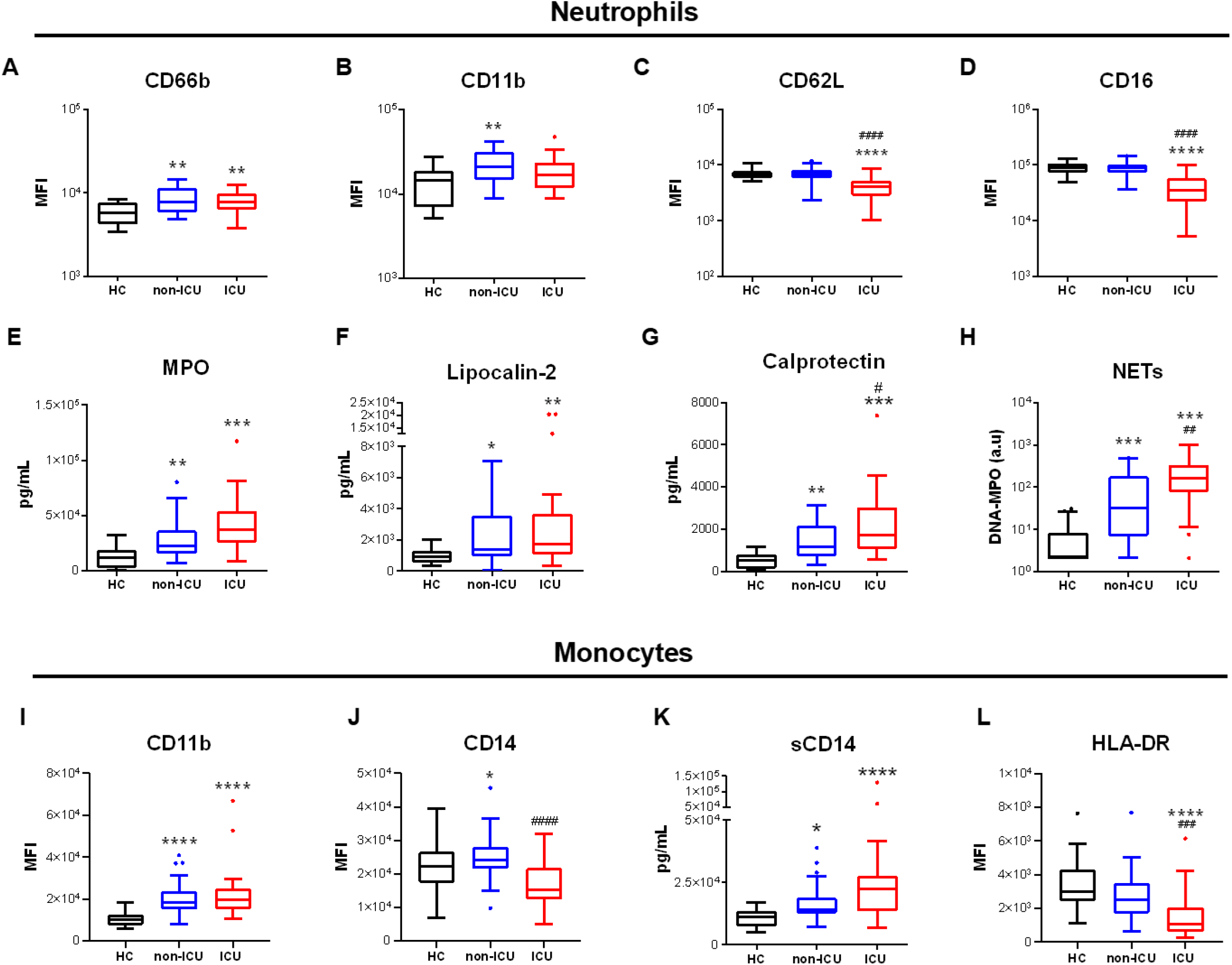
Circulating phagocytes are activated in severe COVID-19. (**A-D**) Expression of activation markers CD66b (A), CD11b (B), CD62L (C), CD16 (D) on neutrophils by flow cytometry. (**E-H**) Concentration in plasma of myeloperoxidase (MPO, E), lipocalin-2 (F), calprotectin (G) and NETs (H) in COVID-19 patients and controls. (**I-J**) Surface expression of CD11b (I) and CD14 (J) on monocytes. (**K**) soluble CD14 concentration in plasma. (**L**) Surface expression of HLA-DR on monocytes. Intergroup comparison by Mann-Whitney U test between healthy controls and ICU or non-ICU patients: ****P < 0.0001, ***P < 0.001, **P < 0.01, *P < 0.05, between non-ICU and ICU patients: ####P < 0.0001, ###P < 0.001, ##P < 0.01, #P<.05. All boxplots whiskers represent 10^th^ and 90^th^ percentiles. HC: healthy controls and degranulation products in the plasma. Moreover, ICU patients exhibit a distinct phenotype with significantly lower CD62L and CD16 expressions on neutrophils, lower CD14 and HLA-DR on monocytes, and higher plasma concentrations of NETs and calprotectin as compared to non-ICU patients.

Monocytes also displayed an activated phenotype with upregulation of CD11b in both patient groups and downregulation of CD14 in ICU patients (Fig.1I-J). Elevated plasma concentrations of sCD14 were seen in both groups (Fig.1K). Interestingly, the activation marker HLA-DR was dramatically downregulated on ICU patients’ monocytes, a feature suggestive of immunoparalysis (Fig.1L).

Altogether, we show that blood monocytes and neutrophils present an activated phenotype in all COVID-19 patients, confirmed by elevated concentrations of circulating activation markers

### Immunosuppressive and immature neutrophil subsets are increased in COVID-19 patients

Activation of innate immunity can give rise to several kind of cell subpopulations with distinct functions. Accordingly, we assessed if phagocyte subpopulation distribution was disturbed in COVID-19 patients. Neutrophil subset identification is a complex and highly debated field since many markers used to define subpopulations are also modulated upon activation*(27)*. However, beside activated cells, several populations of immunosuppressive neutrophils capable of suppressing T cell activation have been described, in particular in cancer*(28)*. Some authors have shown that immunosuppressive or granulocyte Myeloid-Derived Suppressor Cells (MDSC) are characterized by the expression of oxidized lipoprotein receptor LOX-1. Accordingly, we measured expression of LOX-1 and other molecules with immunosuppressive properties (i.e. negative costimulation molecules PD-L1 and ILT3). Percentages of LOX-1+ and PD-L1+ neutrophils were significantly higher in both patient groups (Fig.2A-B) as compared to healthy controls. In contrast, no difference was seen in the percentage of ILT3+ cells between the 3 groups (Fig.2C). Interestingly, however, the percentage of ILT3+ cells was dramatically lower in ICU patients as compared to non-ICU patients and healthy controls in the CD62L^low^ neutrophil subset (Fig. S2).

Beside immunosuppressive neutrophils, an increase in immature neutrophils populations has been described in severe inflammation, in particular in sepsis. Accordingly, we measured surface expression of the maturity markers CD13 and CD10 on neutrophils and showed a dramatic increase of CD13^low^ and CD10^low^ immature neutrophil populations in ICU patients as compared to both non-ICU patients and healthy controls (Fig.2D-E, Fig.S2). Additionally, CD11b^low^CD16^low^ cells, sometimes shown to be banded cells, have been described as another immature neutrophil population *(29)* which we also found to be increased in both non-ICU and ICU patients as compared to healthy controls (Fig.2F).

Several other neutrophil subpopulations have been reported in specific contexts and were also examined in the present study. Pro-angiogenic CD49d+ neutrophils were more frequent in non-ICU patients and even more in ICU patients (Fig.2G) and HLA-DR expression on neutrophils was higher in ICU patients, suggesting an antigen-presenting cell (APC)-like phenotype (Fig.2H). In contrast, CD177+ neutrophils, who do not have clearly defined functions, did not differ between patients and controls (Suppl Fig. 2D). Finally, chemokine receptor expression has also been linked to neutrophil functional properties. In particular, CCR5^hi^ neutrophils have been associated with suppressive functions*(30)*, while CXCR4^hi^ cells, in particular in the CD62L^low^ subset has been linked to “senescent” neutrophils*(31, 32)*. While CCR5 expression was higher in ICU patients, and even in non-ICU patients for the CD62^low^ subset, the expression of CXCR4 did not differ between patients and controls in both CD62L^high^ and CD62L^low^ cells (Fig.S2).

**Figure 2:**
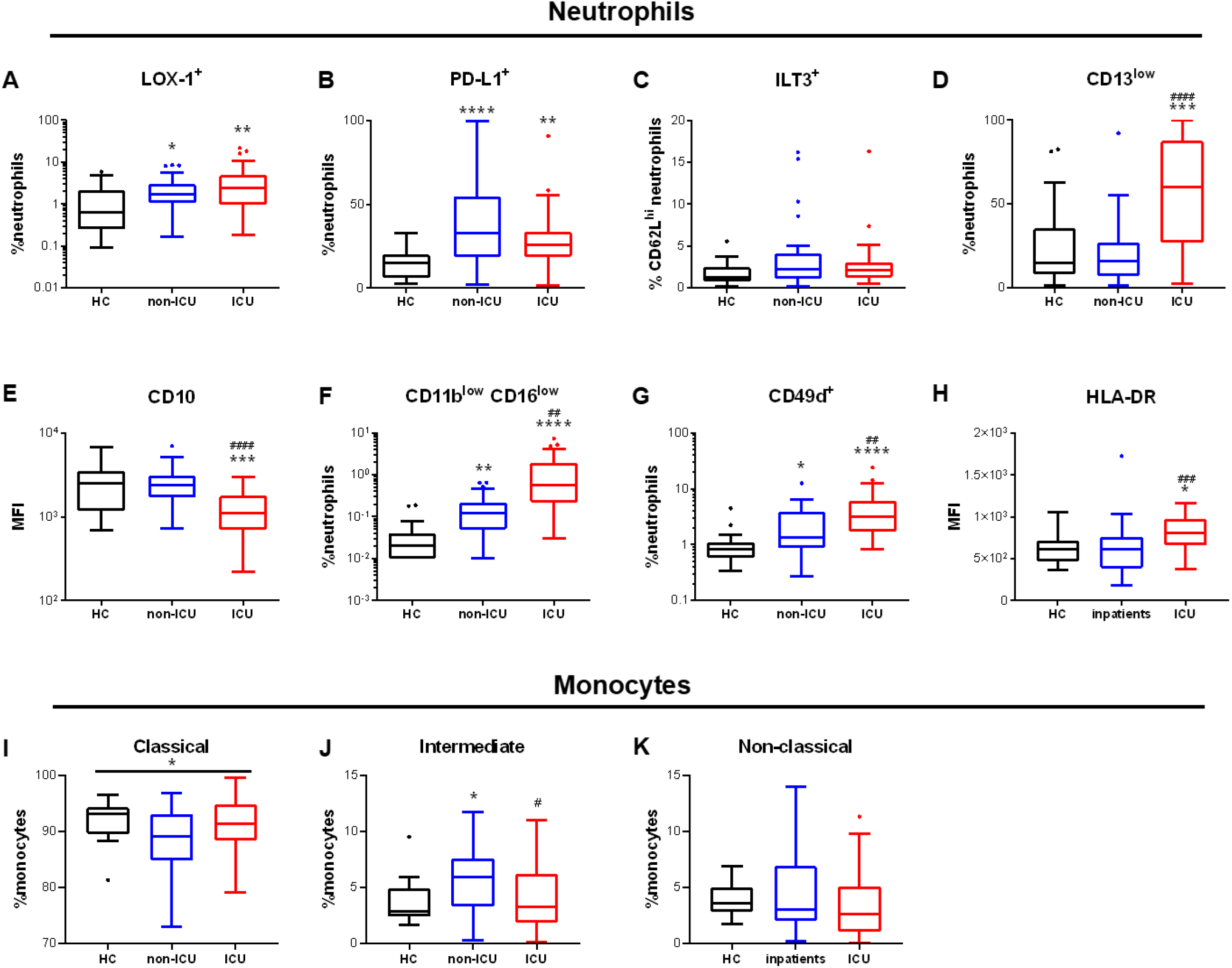
Neutrophil and monocyte subpopulations are disturbed in COVID-19 patients. **(A-H)** Percentages of positive cells and surface expression of markers related to subpopulations in neutrophils by flow cytometry. (A) LOX-1+ neutrophils, (B), PD-L1+ neutrophils, (C) ILT3+CD62L+ neutrophils, (D) CD13^low^ neutrophils, (E) CD10 expression on neutrophils, (F) CD11b^low^CD16^low^ neutrophils, (H) HLA-DR expression on neutrophils. (**I-K**) Percentages of monocytes subpopulations by flow cytometry. (I) CD14+CD16-classical monocytes, (J) CD14+CD16+ intermediate monocytes, (K) CD14^low/-^CD16+ atypical monocytes. Intergroup comparison by Mann-Whitney U test between healthy controls and ICU or non-ICU patients: ****P < 0.0001, ***P < 0.001, **P < 0.01, *P < 0.05, between non-ICU and ICU patients: ####P < 0.0001, ###P < 0.001, ##P < 0.01, #P<.05. All boxplots whiskers represent 10^th^ and 90^th^ percentiles. HC: healthy controls.

In summary, we found a highly disturbed distribution of neutrophil subpopulations in COVID-19 patients. Immunosuppressive LOX-1+ and PD-L1+ neutrophils were increased in both patient groups, while immature neutrophil subpopulations were strongly increased in ICU patients only. As compared to neutrophils, monocyte subpopulations are better described and understood in various disease states. They are classically divided according to CD14 and CD16 expressions in classical (CD14+/CD16-), intermediate (CD14+/CD16+), and non-classical (CD14^low^/CD16+) monocytes. We measured the proportion of these subpopulations and their expression of activation markers. Compared to healthy controls, only intermediate monocytes were significantly higher in non-ICU patients while the proportion of non-classical monocytes was similar in all groups (Fig.2I-K). Moreover, the surface expression of CD11b and HLA-DR was similar in all monocyte subpopulations (Fig. S3), suggesting similar activation among the entire monocyte population. However, CD14 downregulation could be observed only on classical monocytes from ICU patients, while it was significantly upregulated on intermediate monocytes of non-ICU patients. Furthermore, CD62L was significantly upregulated only in atypical monocytes of ICU patients (Fig. S3). These differences suggest that monocytes subpopulations have different responses to the infection and thus may play distinct roles in COVID-19 disease.

All in all, we demonstrate disturbed subpopulation distribution in both neutrophils and monocytes in COVID-19 patients, which was more pronounced in ICU patients. The most striking feature was the increase in immunosuppressive and immature neutrophils in ICU patients, evocative of an impaired immune response. Therefore, we addressed how these phenotypes were associated with patients’ outcome.

### Markers of immature and immunosuppressive phagocytes are associated with severity and poor outcome

To evaluate if the phenotypic changes we observed were linked to disease severity and prognosis, we computed correlations with relevant clinical data. However, ICU and non-ICU patients differ tremendously in terms of severity, demographical data and treatment received (see Table 1). To compensate for such bias, we assessed correlations between phenotype and outcome in non-ICU and ICU patients separately.

**Table 1.**
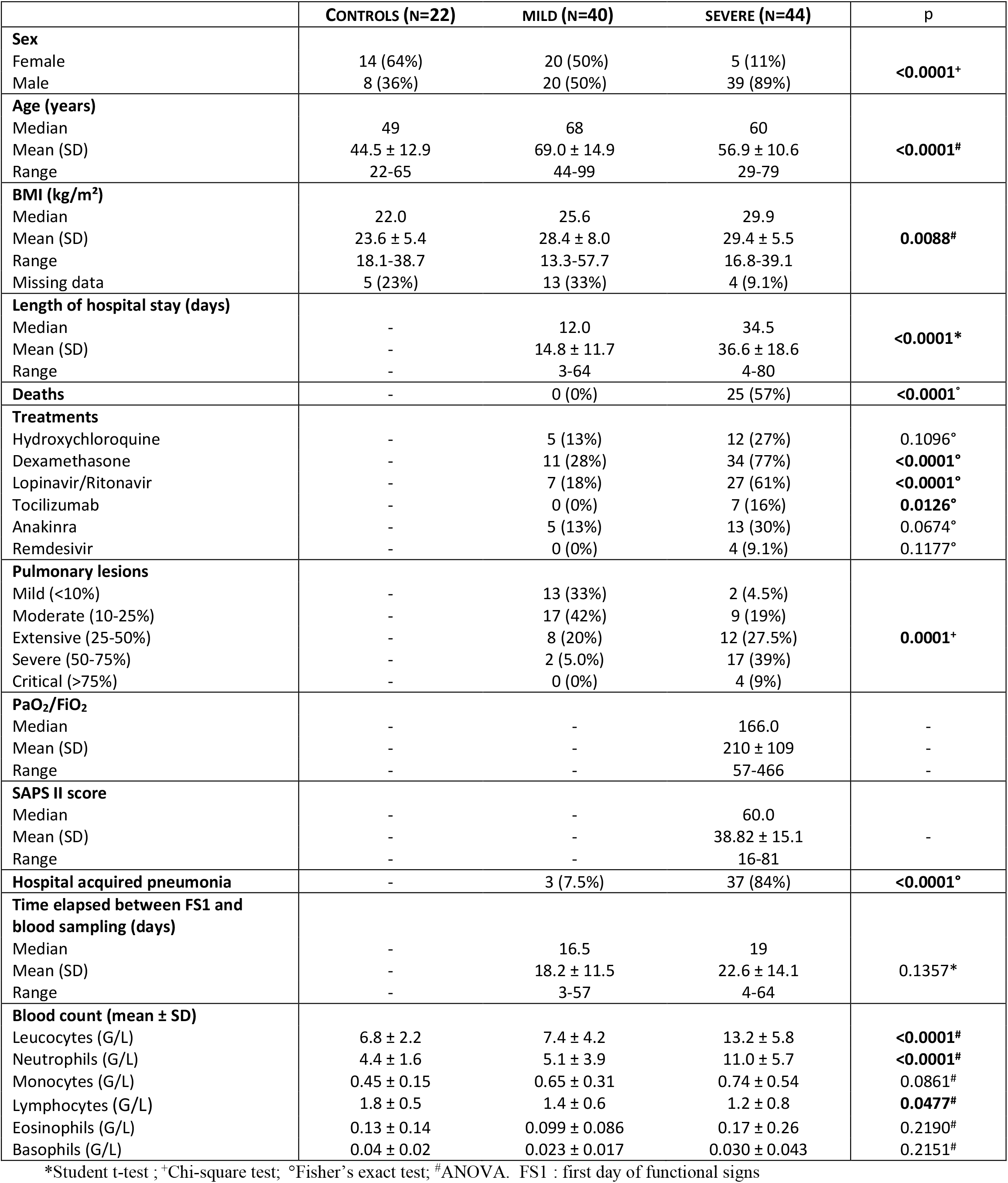
Description of the cohort.

In ICU patients, severity was assessed by the SAPS II organ dysfunction score, the PaO_2_/FiO_2_ ratio, and mortality (Fig. 3A-B). A correlation matrix was performed on main continuous clinical and biological data, and the significant correlations were subjected to hierarchical clustering (Fig.3A). This representation highlights two major clusters of variables correlating together (cluster A and B). The cluster A contains variables correlating with SAPS II score and therefore linked to severity, while cluster B contains the PaO_2_/FiO_2_ ratio and other variables correlating negatively with SAPS II, and therefore linked to less severe disease. Amongst variables linked to severity in those ICU patients, we found several parameters reflecting neutrophil activation, e.g. circulating NETs, elastase, MMP9, and IL-8 plasma concentrations as well as low CD16 expression and increased CD62L^low^ subpopulation. We also found markers associated with immunosuppressive neutrophils like expression of LOX-1, and CCR5. Interestingly, we found a dichotomy between CD62L^high^ (mature/resting) and CD62L^low^ (activated/immature) neutrophils: while higher expression of CD16 and CD10 on CD62L^high^ neutrophils was linked to less severe disease, it was the opposite for the CD62L^low^ population, illustrating that these neutrophil subpopulations have opposite effects on disease outcome. Finally, analysis of monocytes showed mostly that severity was linked with lower CD14 and HLA-DR expression on classical monocytes.

**Figure 3:**
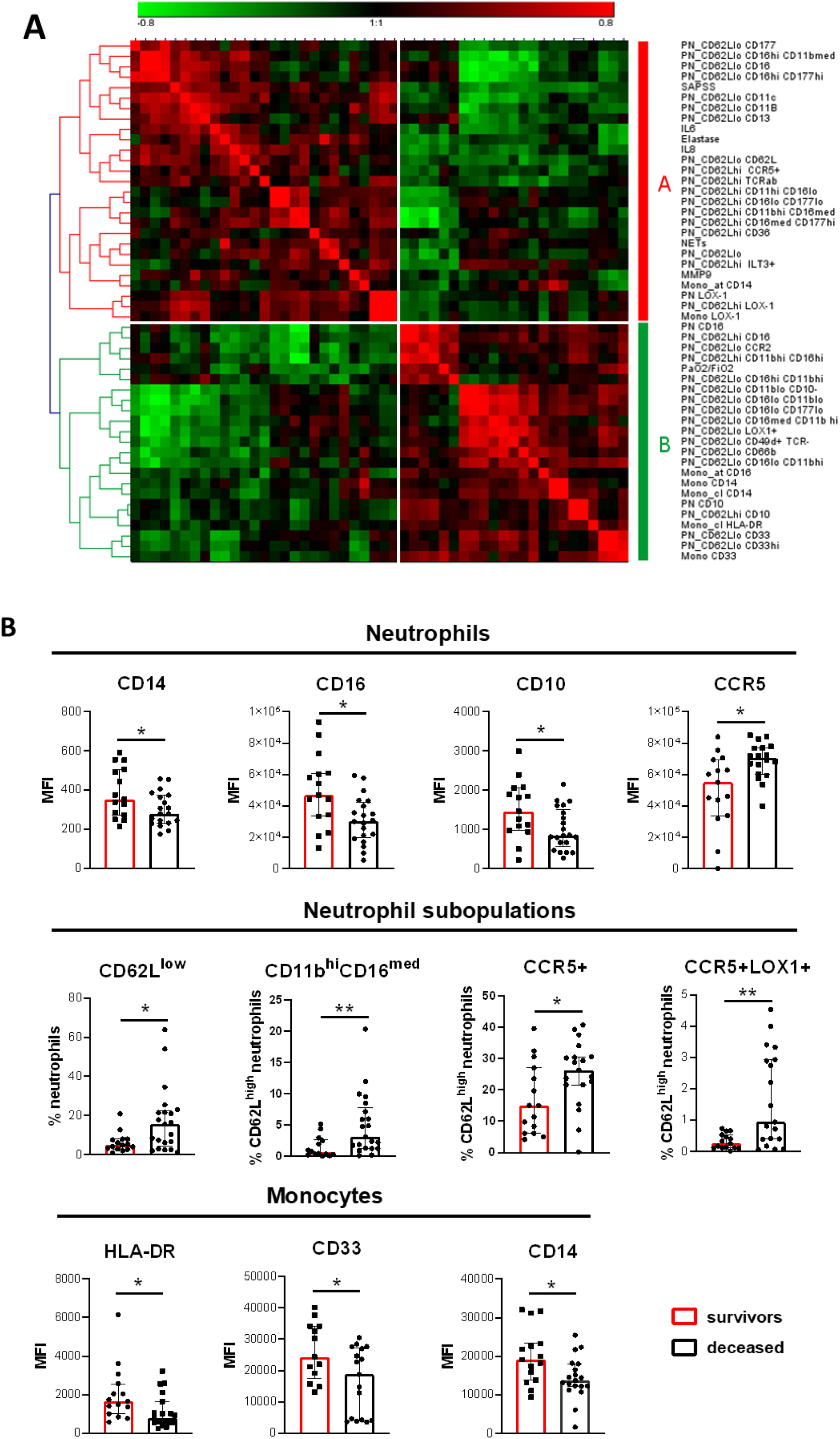
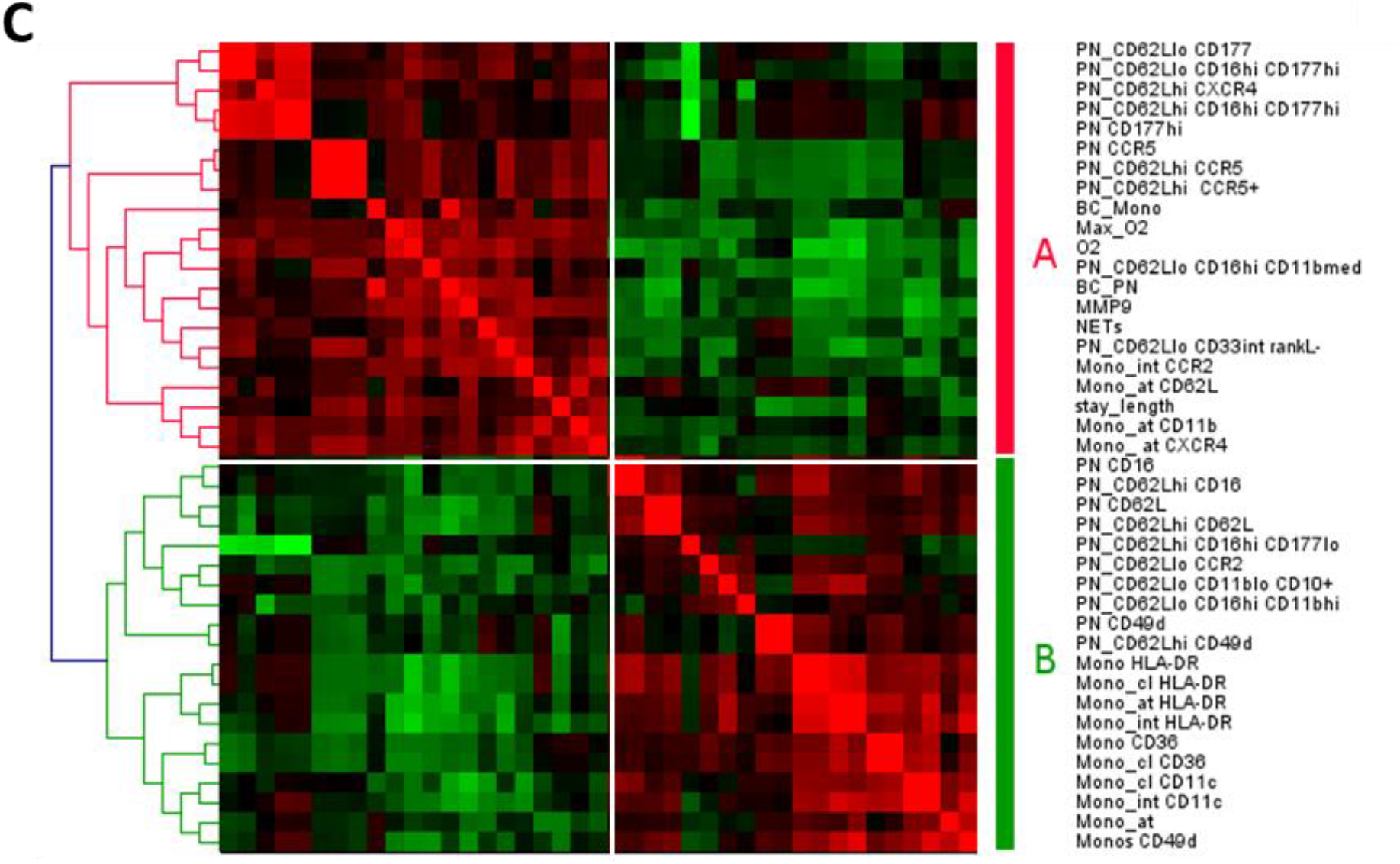
Association of innate immunity-related markers with severity. **(A**) Hierarchical clustering of the phenotypic and soluble markers significantly correlated with SAPS II score or the PaO_2_/FiO_2_ ratio in ICU patients. Red is for positive correlation and green for negative correlation. Intensity of color is proportional to Spearman correlation coefficient. Cluster A groups markers with a correlation profile similar to SAPS II score. Cluster B groups markers with correlation profiles similar with PaO_2_/FiO_2_ ratio. (**B**) Markers significantly different between patient who survived (red) or died (black) during their ICU stay. The histograms represent the median and the error bars the interquartile range. Intergroup comparison by Mann-Whitney U test between survivors and deceased patients, **P < 0.01, *P < 0.05. (**C**) Hierarchical clustering of the phenotypic and soluble markers significantly correlated with external oxygen requirement on the day of sampling (O_2_), maximal external oxygen requirement (Max_O2) or length of hospitalization in non-ICU patients (stay_lengh). Red is for positive correlation and green for negative correlation. Intensity of color is proportional to Spearman correlation coefficient. Cluster A groups markers with a correlation profile similar to O2, Max_O2 and stay_length. Cluster B groups markers negatively correlated to cluster A. BC_PN: neutrophil absolute count; BC_Mono: monocyte absolute count.

**Figure 4:**
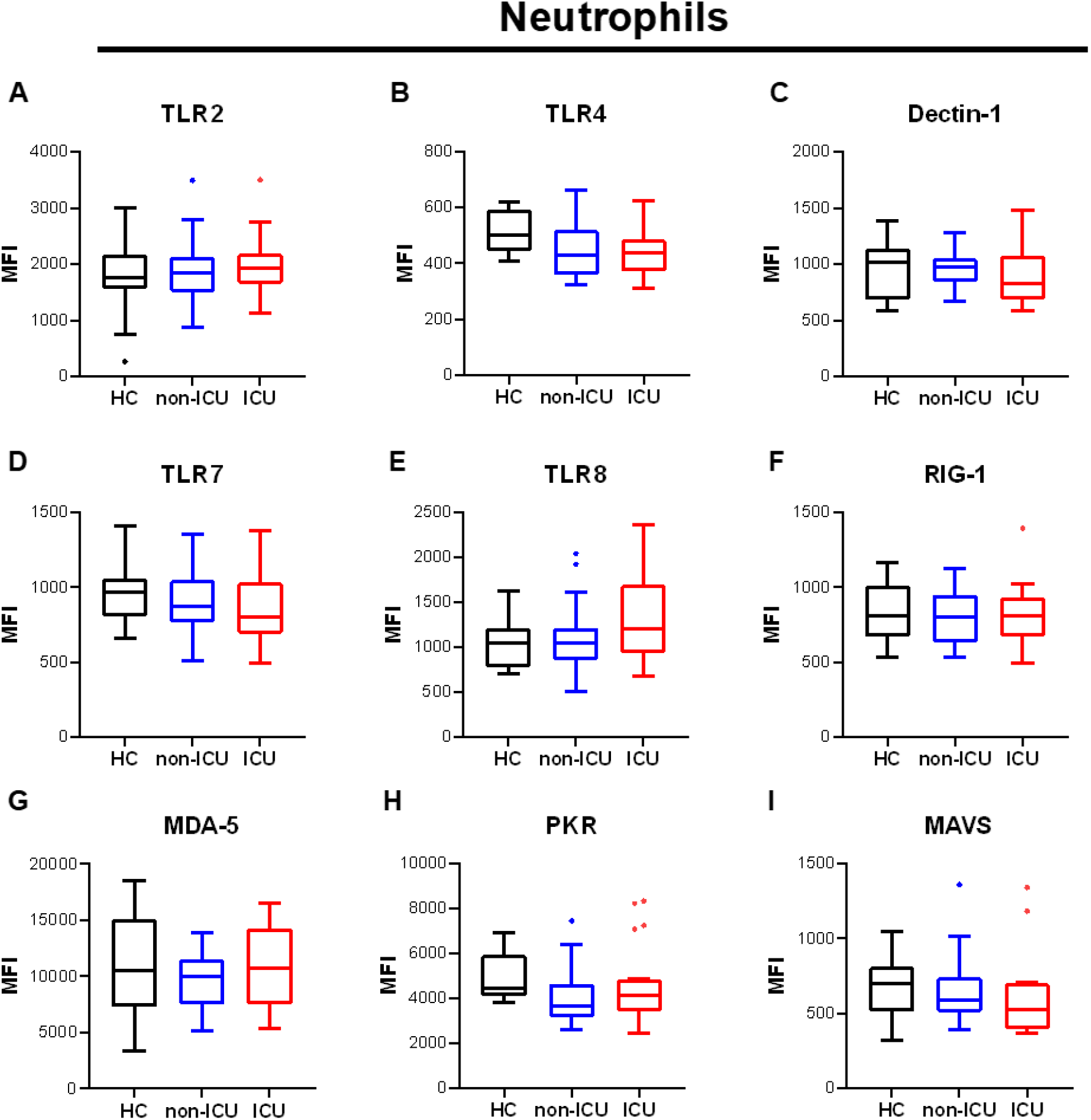
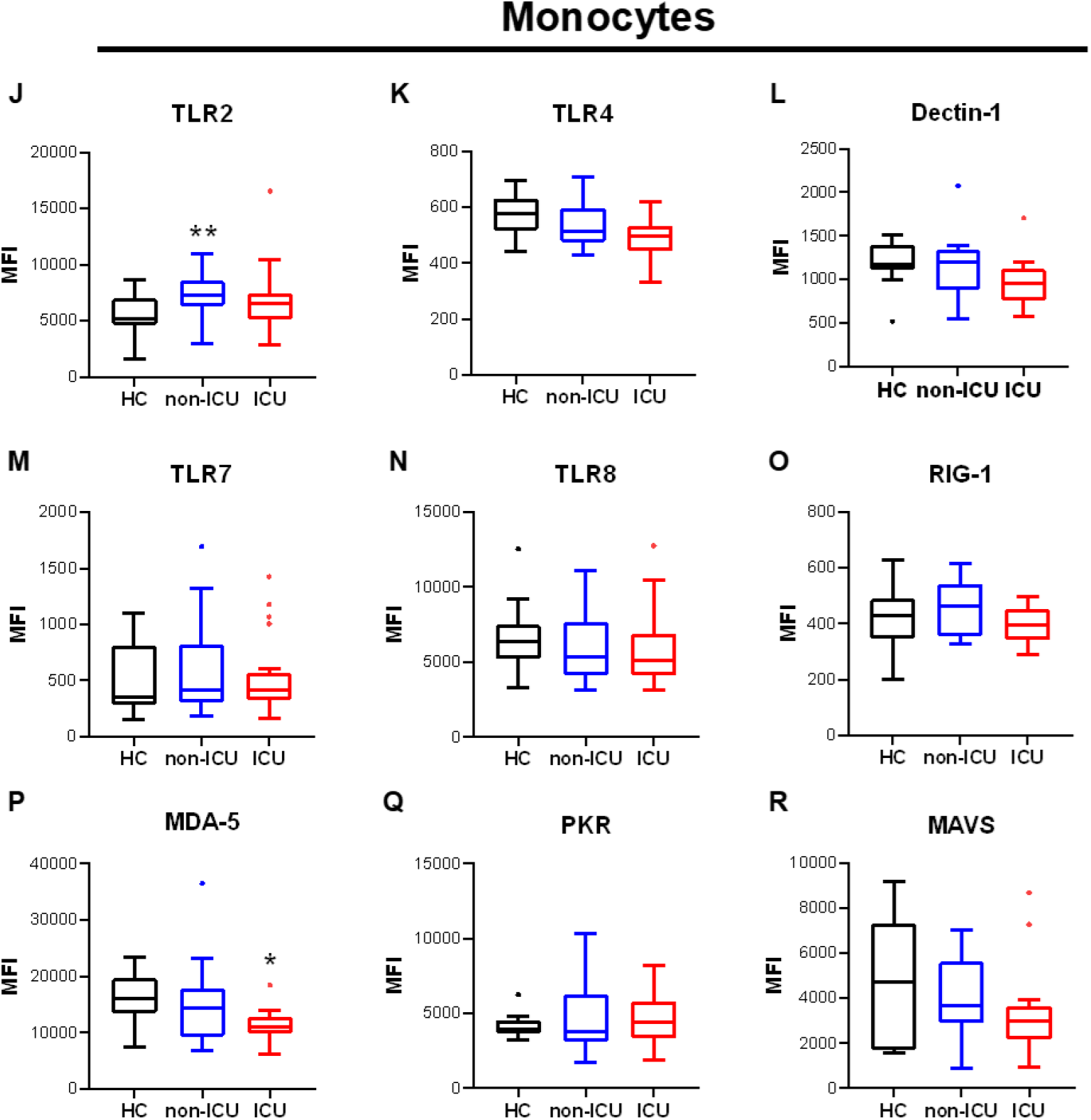
Expression of pattern-recognition receptors on circulating phagocytes. **(A-C)** Surface expression of plasma membrane pattern-recognition receptors (PRR) on neutrophils: (A) Toll-like receptor 2 (TLR2), (B) TLR4, (C) Dectin-1. (**D-E**): intracellular expression of endosomal PRR on neutrophils: (D) TLR7, (E) TLR8. (**F-H**): Intracellular expression of cytosolic PRR in neutrophils: (F) RNA-sensing retinoic acid-inducible gene I (RIG-1), (G) melanoma differentiation-associated gene-5 (MDA-5), (H) protein kinase R (PKR). (**I**) Intracellular expression of Mitochondrial Antiviral Signaling Protein (MAVS). **(J-L)** Surface expression of PRR on monocytes: (J) TLR2, (K) TLR4, (L) Dectin-1. (**M-N**): intracellular expression of endosomal PRR on monocytes: (M) TLR7, (N) TLR8. (**O-Q**): Intracellular expression of cytosolic PRR in monocytes: (O) RIG-1, (P) MDA-5, (Q) PKR. (**R**) Intracellular expression of MAVS in monocytes. Intergroup comparison by Mann-Whitney U test between healthy controls and ICU or non-ICU patients, **P < 0.01, *P < 0.05. All boxplots whiskers represent 10^th^ and 90^th^ percentiles.

**Figure 5:**
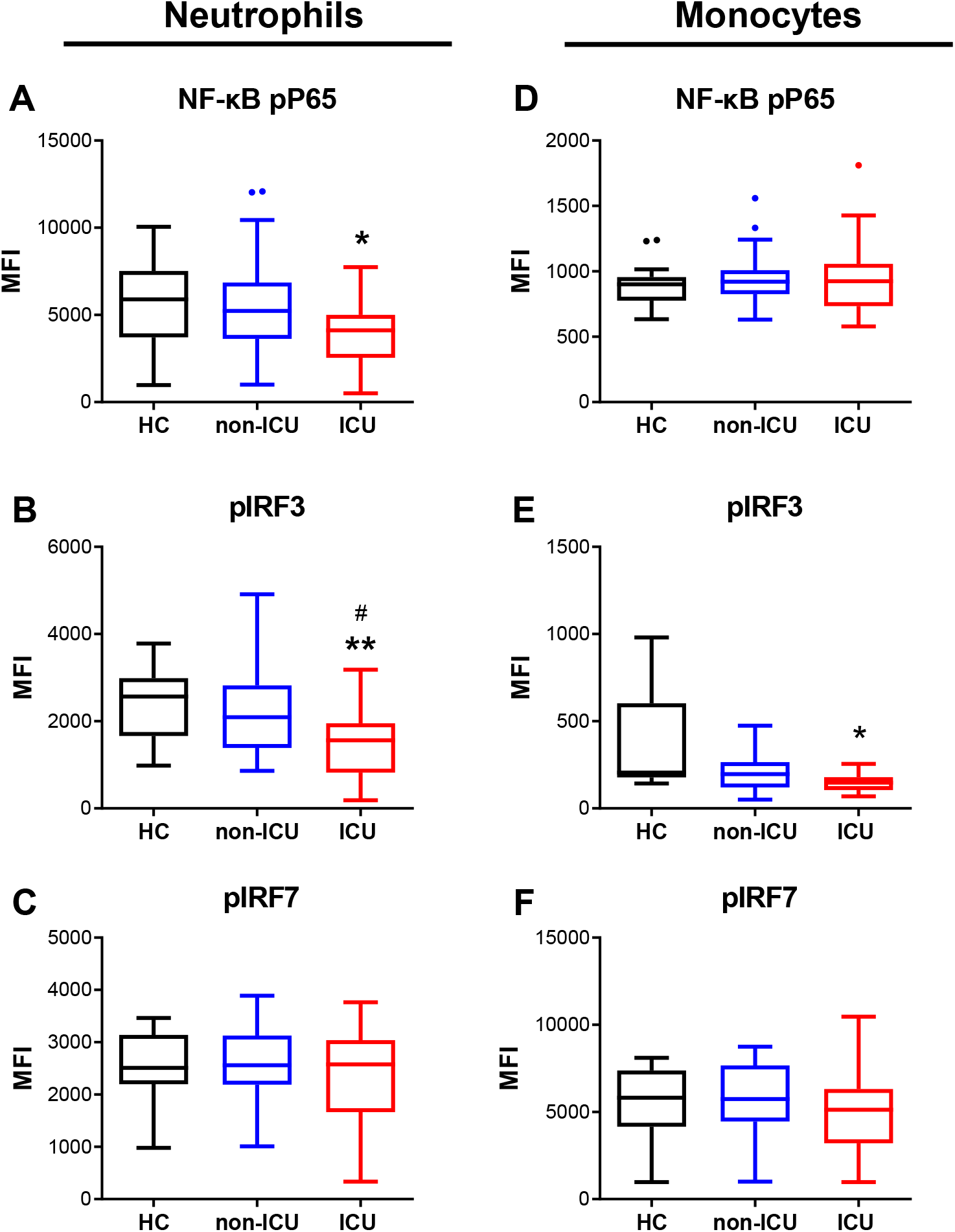
Pattern-recognition related signaling pathways activation in circulating phagocytes. Intracellular expression of phosphorylated signaling molecules from PRR pathways in (**A-C**) neutrophils and (**D-F**) monocytes by flow cytometry. (A,D) Expression of phosphorylated Interferon Response Element 3 (IRF-3), (B,E) expression of phosphorylated IRF-7, (C,F) expression of phosphorylated Nuclear Factor B protein p65 (pp65). Intergroup comparison by Mann-Whitney U test between healthy controls and ICU or non-ICU patients, **P < 0.01, *P < 0.05. All boxplots whiskers represent 10^th^ and 90^th^ percentiles

We then assessed if phagocyte phenotype was linked to mortality by comparing the expression of markers between survivors and deceased ICU patients (Fig. 3B, Figs. S4-S5).

Neutrophils of deceased patients had significantly lower CD16 and CD10 expression which corresponded to more activated and less mature cells. The CD62L^low^ and CD11b^hi^ CD16^med^ activated neutrophil subpopulations were also increased. Additionally, we found an increased expression of CCR5, and in particular of the CCR5+LOX-1+ subpopulation, which has been described as immunosuppressive. Monocytes from deceased patients had lower expression of HLA-DR, CD33 and CD14. A lower HLA-DR expression could be seen in all monocyte subpopulations; however, the difference between survivors and deceased patients was most striking in intermediate monocytes. Moreover, non-classical monocyte subpopulation had a distinct phenotype in deceased patients, with higher expression of CD62L and CD11b but lower expression of CD11c and HLA-DR (Fig. S4). Finally, the analysis of soluble parameters revealed that circulating concentrations of IL-6 and neutrophil elastase were significantly higher in deceased patients (Fig. S5).

Interestingly, we found 6 parameters in common between cluster analysis and the markers associated with lethal outcome, suggesting they have robust link with severity. These markers were a higher expression of CCR5 and LOX-1, a lower expression of CD16, CD10, and CD62L on neutrophils, and a lower expression of CD14 and HLA-DR on monocytes.

In the group of non-ICU patients, no unfavorable outcome was recorded during the study time (i.e., no death or ICU admission). We therefore assessed severity by exogenous oxygen (O_2_) requirement and duration of stay in the hospital and performed the same analysis than for ICU patients (Fig.3C). As for ICU patients, this analysis allowed to identify two main clusters: one cluster of variables correlated with O_2_ requirement and duration of stay and was therefore associated with severe disease (A), and the other cluster of variables negatively associated with cluster A and therefore with less severe disease (B). Variable analysis showed that neutrophil activation markers like circulating MMP9 and NETs, and low expression of CD16 and CD62L on neutrophils were linked to severe disease. This was also the case for high expression of CCR5 and CXCR4 and low expression of CD49d. Contrarily to ICU patients, many monocyte-related variables were highlighted in this analysis, in particular association of severity with lower expression of HLA-DR on all monocyte subpopulations, as well as lower expression of CD36, CD11c, and CD49d. As for ICU patients, atypical monocytes displayed specific associations with higher expressions of CD62L, CD11b or CXCR4 associated with severity.

Globally, our analyses show many phagocyte-related features significantly associated with disease severity in both ICU and non-ICU patients. Some of these features were common to both groups of patients: severity was linked to lower CD16 and CD62L expression and higher CCR5 expression on neutrophils, lower HLA-DR expression on monocytes, and higher concentrations of MMP9 and NETs. Additionally, some other features were found only in one patient group like the deleterious effect of higher LOX-1 and lower CD10 in ICU patients, highlighting the role of immunosuppressive and immature neutrophils in this group of patients. In contrast, we observed a protective effect of neutrophil CD49d and monocyte CD36 only in non-ICU patients.

Since some features were associated with activated phagocytes, but other to suppressive or immature and therefore less active cells, we next assessed what was the resulting effect of these phenotypical features on phagocyte functions.

### Phagocytes of COVID-19 patients display impaired anti-microbial functions

The first step in innate immune response is danger recognition by pattern-recognition receptors (PRR) expressed on myeloid cells. We therefore analyzed the expression of the main extracellular (TLR2, TLR4, Dectin-1), endosomal (TLR7, TLR8) and cytoplasmic (RIG-1, MDA-5) PRRs as well as two major adaptor proteins of cytoplasmic PRRs, (PKR and MAVS) on neutrophils and monocytes (Fig.4). PRR expression on phagocytes was globally similar in patients and in controls (Fig.4A-R), except for TRL2 that was significantly increased in monocytes of non-ICU patients (Fig.4J) and the cytosolic sensor MDA-5 that was slightly decreased in ICU patients (Fig.4P). These slight variations, however, seem unlikely to result in a major effect in pathogen response. To confirm this result and explore further pathogen-sensing functions, we looked at the levels of active phosphorylated signaling molecules downstream of PRR activation, namely pNF-κB (p65), pIRF-3 and pIRF-7 (Fig.5). Interestingly, intracellular expression of pIRF3 and pNF-κB were significantly lower in neutrophils from ICU patients (Fig.5A-B) while expression of pIRF3 was lower in monocytes (Fig.5E), suggesting that phagocytes are less responsive in these patients, which could have an impact on their sensitivity to pathogens.

We next tested phagocytosis capacity using pHRodo-conjugated zymosan particles. We show that phagocytosis was significantly impaired in both neutrophils and monocytes from ICU and non-ICU patients (Fig 6A-B). These cells have thus a lower ability to clear pathogens, which was not restricted to ICU patients but also occurred to a lesser extent in non-ICU patients.

**Figure 6:**
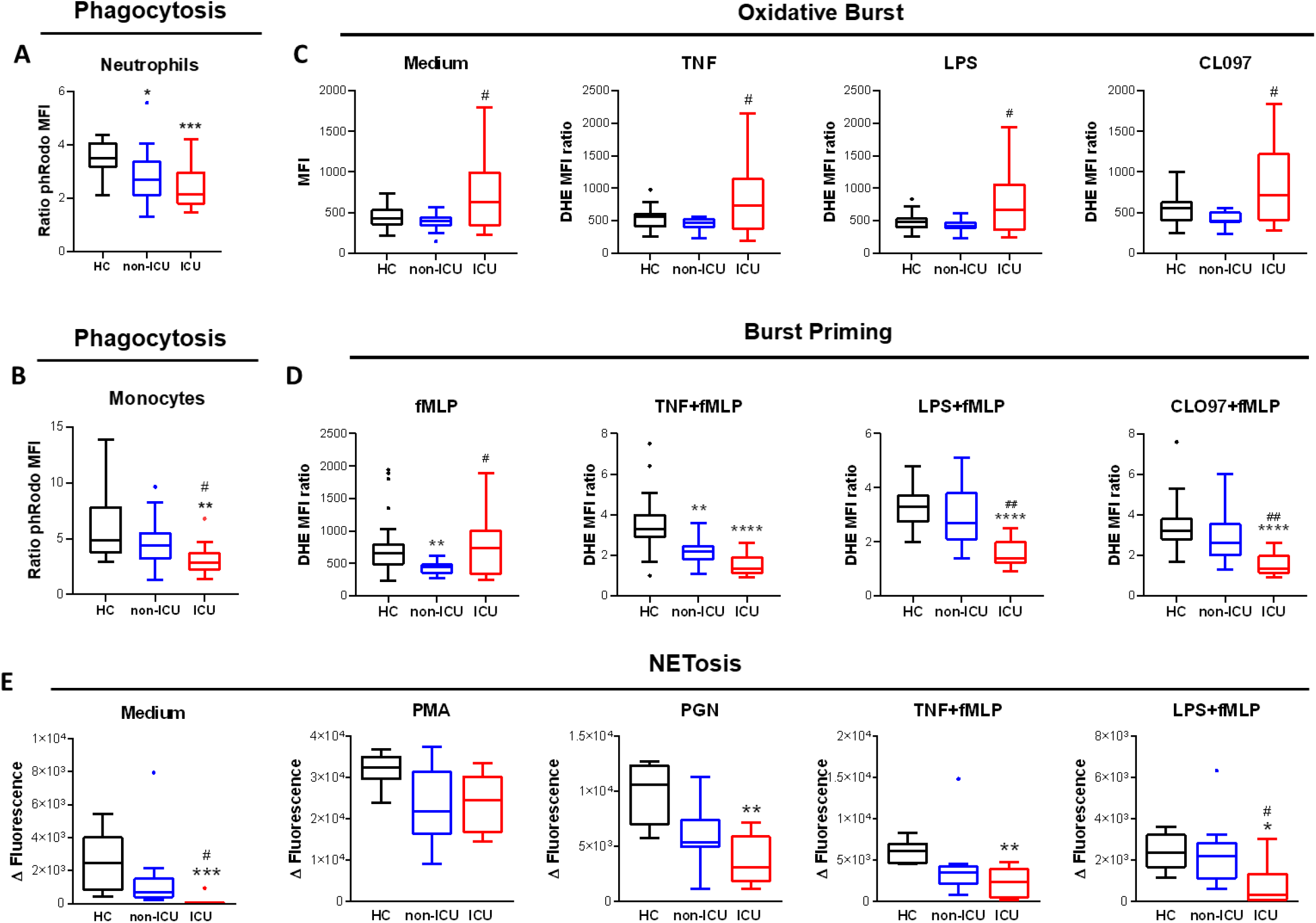
Anti-microbial functions of circulating phagocytes are impaired in severe COVID-19. **(A-B)** Measurement of phagocytosis capacity by phRodo-conjugated Zymosan particles uptake in (A) neutrophils and monocytes. (**C-D**) Measurement of (C) oxidative burst in response to medium, Tumor Necrosis Factor alpha (TNF), lipopolysaccharide (LPS), TLR8 ligand CLO97, and (B) priming of formyl-methionine-leucine-phenylalanine (fMLP)-induced burst by TNF, LPS and CLO-97 in neutrophils. (**E**) Measurement of neutrophils NETosis capacity in response to medium, phorbol myristate acetate (PMA), peptidoglycan (PGN), TNF and fMLP, LPS and fMLP. Intergroup comparison by Mann-Whitney U test between healthy controls and ICU or non-ICU patients, ****P < 0.0001, ***P < 0.001, **P < 0.01, *P < 0.05., between non-ICU and ICU patients, ##P < 0.01, #P<.05. All boxplots whiskers represent 10^th^ and 90^th^ percentiles.

Neutrophil oxidative burst is a critical mechanism of host defense and patients with impaired oxidative burst suffer from severe recurrent infections. Neutrophils from ICU patients showed higher reactive oxygen species (ROS) production as steady state that non-ICU patients and healthy controls. This production did not change in response to priming agents (TNF, LPS, TLR8 agonist CLO97) and weak burst agonist formyl-methionine-leucin-phenylalanine (fMLP, Fig.6C). Additionally, burst priming by combining fMLP with priming agents was significantly lower in ICU patients, and even in non-ICU patients for the TNFα priming condition (Fig.6D). Altogether, these results suggest that neutrophils are hyperactivated at basal state in ICU patients but exhibit a decreased capacity to be reactivated, suggesting higher susceptibility to bacterial or fungal infections.

The production of NETs, or NETosis, has been shown to be critical for the control of some infections, in particular for filamental fungus like *Aspergillus sp* infections that were described in severe COVID patients. Accordingly, we measured the NETosis capacity of freshly isolated neutrophils from COVID-19 patients (Fig.6E). In response to PMA, a strong pharmacological trigger, NET release was similar in patients and controls. However, spontaneous NETosis (in the absence of stimulus) and NETosis in response to various physiological ligands like TLR2 ligand peptidoglycan (PGN) or fMLP combined with TNFα or LPS was significantly decreased in ICU patients.

Altogether, these functional results strongly suggest that neutrophils and monocytes of severe COVID-19 patients are dysfunctional and exhibit a decreased capacity to fight against pathogens, whatever the function tested (PRR sensing, phagocytosis, oxidative burst, NETosis). Such impaired functions are likely to have an impact on host immune response and thus be deleterious for the patient outcome. Therefore, we measured the association of phagocyte functionality with disease severity and prognosis.

### Neutrophil and monocyte dysfunctions are associated with poor prognosis in ICU patients

To quantify neutrophil and monocyte antibacterial functions we chose phagocytosis as an indicator since it involves several cellular mechanisms (receptor recognition, particle engulfment, and vacuole acidification) and was decreased in both patient groups (Fig.7).

**Figure 7:**
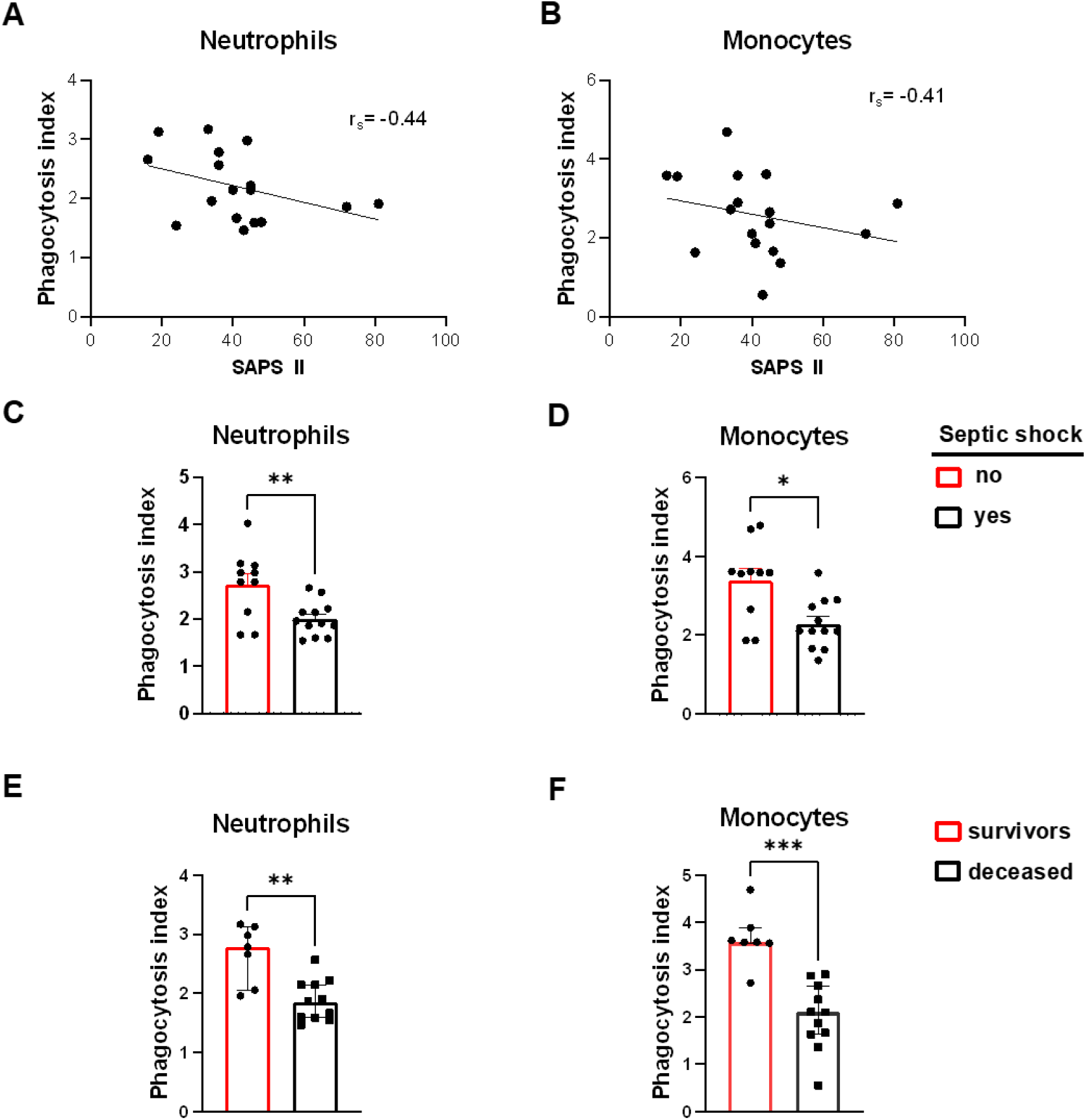
Functional impairment of phagocytes is linked to organ dysfunction, septic shock, and mortality in ICU patients. **(A-B)** Correlation between phagocytosis and SAPS II in (A) neutrophils and (B) monocytes. Rs: Spearman correlation coefficient Rho. **(C-D)** Phagocytosis capacity in patients with (black) or without (red) a septic shock of (C) neutrophils and (D) monocytes. The histograms represent the median and the error bars the interquartile range. Intergroup comparison by Mann-Whitney U test, **P < 0.01, *P<0.05. **(E-F)** Phagocytosis capacity in survivors (red) and deceased (black) ICU patients of (E) neutrophils and (F) monocytes. Intergroup comparison by Mann-Whitney U, ***P < 0.001, **P < 0.01.

Phagocytosis impairment was linked to organ dysfunction as it correlated negatively with SAPS II score for both cell types (Fig.7A-B). Additionally, patients who presented with septic shock had significantly less phagocytic activity that patients that did not (Fig.7C-D). Finally, phagocytosis capacity was much lower in patients who died as compared with patients who survived. (Fig.7E-F).

We thus conclude that neutrophil and monocyte functionnal impairment in COVID-19 patients are linked to severity and poor income, at least in part by favoring the occurrence of severe infections.

## DISCUSSION

In this study, we investigated the links between circulating innate phagocyte phenotype and functions and severity in COVID-19 patients. We found cellular and molecular features of myeloid cell activation and disturbed subpopulation distribution in both ICU and non-ICU patients. However, ICU patients presented a distinct set of parameters suggestive of immunosuppressive and immature neutrophils that clearly distinguished them from non-ICU patients. NETs and MMP-9 plasma concentrations, overexpression of CCR5 and downregulation of CD16 and CD62L on neutrophils, as well as downregulation of HLA-DR on monocytes were associated with severity and prognosis in both non-ICU and ICU patients. Overexpression of LOX-1 and downregulation of CD10 on neutrophils, were linked to severity in ICU patients only, while overexpression of CXCR4 and downregulation of CD49d on both cell types was detrimental in non-ICU patients. Most importantly, we show by exploring several cellular functions that neutrophils and monocytes from both patient groups had impaired antimicrobial mechanisms and that this loss of function was linked to organ dysfunction, severe infections, and mortality.

A detrimental role for activated myeloid cells has long been documented in various adult lung diseases, in particular in ARDS *(33, 34)*. In COVID-19, multiple clues point to a major contribution of the innate response in the disease pathophysiology. In ICU patients, neutrophils and NETs have been shown to infiltrate lungs *(35)*, while transcriptomic studies showed a link between death and neutrophil activation *(36)*, and monocyte expression of proliferation marker Ki-67 was linked to severity *(37)*. Moreover, in a mouse model of SARS-Cov2 infection, neutrophils with an aberrant phenotype accumulated in the lungs *(38)*. In line with these results, we found activated neutrophils and monocytes in the blood not only in ICU patients but also in non-ICU patients, implicating that myeloid cells are involved even in mild disease. This raises the issue of circulating phagocytes activation mechanism. SARS-CoV2 virus is detected mostly in patients’ respiratory tract but not in the blood stream except at low levels in critically ill patients and is thus an unlikely candidate for circulating immune cell activation. It seems more likely that a yet undefined combination of circulating pro-inflammatory factors from infection sites could be involved. In critical patients, many inflammatory cytokines have been measured in the bloodstream, some of which could play this activation role. However, elevated cytokine concentrations are not found in most non-ICU patients. An interesting candidate could be an epithelial-secreted alarmin, as it was shown in animal models that SARS-Cov2 infection leads to a robust secretion of S100A8/A9 protein by the airway epithelium that is critical to the pathogenesis *(38)*.

We found several circulating and membrane activation markers of neutrophils linked to severity and prognosis in COVID-19 patients. This is in line with large-scale studies that have correlated basic immune parameters and outcome in COVID-19 patients and reproductively found association of neutrophilia and/or neutrophil-to-lymphocyte ratio with severity *(12, 39)*. A few reports used more detailed phenotyping and found activation markers and myeloid subpopulations linked to prognosis, some in agreement with ours (NETs *(17, 40)*, elastase *(40)*, MMP9 and calprotectin *(41)*, HLA-DR^low^ monocytes *(42, 43)*, PD-L1^+^ neutrophils *(44, 45)*, CD10^low^ neutrophils *(43)*) and some that we did not reproduce (CD14^hi^ monocytes*(46)*, lactoferrin*(41)*). The discrepancies between the studies are likely due to differences in methodologies used (cytometry, RNAseq) and/or in the populations selected (all type of patients or severe only). Most importantly however, none of these studies separated ICU from non-ICU patients, which is ground for numerous biases since both patient populations significantly differ for sex, age, and BMI, and some ICU-specific treatments like mechanical ventilation are known to activate neutrophils *(47)*. Here we calculated correlations between immune and clinical parameters in each patient group separately. Interestingly, some parameters were associated with severity in both patient groups, most of them being neutrophil activation markers (e.g. NET or MMP9 concentrations, or downregulation of surface CD62L and CD16), which advocates for a globally deleterious role of neutrophils in both ICU and non-ICU patients. Other markers, like CCR5 overexpression, have been shown to be associated with immunosuppressive neutrophils and lower survival in several cancer reports *(48)*, which could imply that these cell play a role in suppression of adaptive immune response in COVID an explain their link with poorer prognosis. In the same line of thought, HLA-DR downregulation on monocytes is a marker known to be linked to immunoparalysis and severity in sepsis *(49, 50)*. Variables linked to severity in ICU patients included increased expression of LOX-1, downregulation of CD10 and elevated neutrophil elastase concentrations. LOX-1 is a receptor to oxidized lipids and has been shown to play a role in neutrophil recruitment to the lungs *(51)*, but also identifies suppressor neutrophils in cancer *(52)*, two features susceptible to play a major role in COVID pathophysiology. CD10 is a maturity marker, and low CD10 expression has been associated with immature and less functional neutrophils *(53, 54)*. Finally, neutrophil elastase enzymatic activity is strongly suspected to participate in lung injury during ARDS *(55)*. Thus, compared with non-ICU patients, ICU patients present additional features of proinflammatory immune components but also immature and immunosuppressive cells that contribute to global immune dysfunction and may explain their more severe condition.

To understand the role of blood myeloid cells in COVID-19 pathogenesis, it is important to evaluate not only the populations involved but also their functionality. We tested four typical myeloid cell functions (PRR signaling, phagocytosis, oxidative burst, NETosis), and found that cells from COVID patients were less functional than cells from healthy controls. This was much more pronounced in ICU patients, even if some level of dysfunction could be seen in non-ICU patients. While this may seem paradoxical in regard to the elevated expression of activation markers that we and others have described, this is similar to the immunoparalysis setting described in severe sepsis *(56)*. In this setting, neutrophils have defective oxidative burst and phagocytosis despite extremely high expression of cellular and soluble activation markers *(56, 57)*. Innate immunity is often described as a double-edged sword, both beneficial and harmful though anti-microbial and pro-inflammatory properties. Our findings suggest that in severe COVID-19, not only neutrophil and monocyte activation by-products are deleterious, but also that immune response is impaired through immature and immunosuppressive myeloid cell populations. The links we found between innate immune dysfunction, disease severity and mortality strongly suggest a pivotal role for innate phagocytes in loss of control of the infection and ultimately, in disease outcome.

There are some limitations to this study. First, this study was performed in the middle of the first wave of the COVID-19 pandemic in France, when many treatments were tested in parallel on critical patients, increasing variability between patients and reducing our ability to detect small effects. In the same line of thought, immunomodulatory treatments were used either on almost all patients (e.g. corticosteroids) or in a small number of patients for each molecules (e.g. tocilizumab), which precludes from in-depth multivariate statistical analysis that could control for their use. Finally, it was not possible for logistic reasons to realize a longitudinal sampling of the patients, which prevent us to reconstitute the precise trajectory of immune responses.

Despite these limitations, we were able to see some strongly significant associations that we are in the process of confirming on a larger cohort from the second and third wave of infections.

The fact that neutrophil activation parameters were associated with poor prognosis in both patient groups is a strong incentive for neutrophil-targeting clinical trials, some of which are already ongoing. For example, N-acetylcysteine has been proposed as an antioxidant treatment but a first phase III clinical trial was negative *(58)*. The use of intratracheal DNAse (dornase alfa) to dismantle NETs has been tried in small cohorts and at least one clinical trial is ongoing. Finally, use of a potent elastase inhibitor, sivelestat has been suggested but remains to be tested *(59)*. It must be stressed that all these approaches will also affect anti-microbial functions of myeloid cells. As we found that these cells were already dysfunctional and since control of secondary infections is crucial in critical patients, we greatly caution that future investigators should take into account the balance between control of inflammation and anti-microbial activity. A specific time-window for anti-cytokine efficacy in COVID patients has been suggested *(60)*. Since we found immunosuppressive cells and loss of function mostly in ICU patients, we hypothesize that suppressive therapies targeting neutrophils or monocytes would be more indicated in at-risk non-ICU patients to prevent progression to critical stage, while in ICU patients strategies aiming at stimulating anti-microbial properties of immune cells would be more appropriate.

In conclusion, we found that innate immune deficiency is present in both non-ICU and ICU COVID-19 patients and associated with disease severity and prognosis. The precise trajectories of myeloid subpopulations and functions according to disease stage should be further investigated and taken in account for future treatment options.

## MATERIALS AND METHODS

### Study design

A total of 84 consecutive Covid-19 patients from Bichat Hospital, Paris, France, were included during the first epidemic wave (March-June 2020). Patients were hospitalized in a standard hospital ward in the Infectious Disease Department (non-ICU, n=40) or in intensive care unit for critically ill patients (ICU patients n=44). Demographic, clinical, and biological data of patients are summarized in Table 1. Twenty-two healthy blood donors were added to establish the normal range of studied parameters. The study was approved by National Ethics committee CEEI/IRB under the number 20-715.

### Whole blood myeloid cell phenotyping

EDTA-treated blood samples were incubated with several antibody combinations containing a wide selection of markers for neutrophil and monocyte subpopulations and activation (see Supplemental Table 1 for antibody list). After 30 min at 4°C, samples were subjected to red blood cell (RBC) lysis (FACS Lysing solution, BD BioSciences) for 10 min and washed before acquisition. Intracellular targets were stained using the CytoFix/CytoPerm kit (BD Biosciences) according to manufacturer’ s instructions. All samples were acquired on a FACS Lyrics cytometer (BD Biosciences), and analysis of data was done on FlowJo 10.0 (this applies to all flow cytometry experiments of this study).

### Signaling pathways analysis

To document the signaling pathways involved in phagocyte activation by pattern-recognition receptors (PRR), flow cytometric analysis of phospho-NF-κB (p65), phospho-IRF7, and phospho-IRF3, was performed using BD Phosphoflow reagents (BD Biosciences), according to manufacturer’ s instructions. Briefly, 200µL of EDTA-treated blood was incubated with anti-CD14 for 15min at room temperature, then blood cells were lysed and fixed using pre-warmed BD Phosphoflow Lyse/Fix Buffer for 10min at 37°C. After washing in PBS, cells were permeabilized using BD Phosphoflow Perm Buffer III and chilled 30min on ice. Cells were then washed twice in PBS and stained intracellularly using appropriate antibodies (see Suppl Table 1). After 30min of incubation at room temperature in the dark, cells were washed again and finally acquired on a Lyrics flow cytometer.

### Phagocytosis assay

Heparinized whole blood was incubated with pH-sensitive pHRodo-conjugated Zymosan bioparticles (ThermoFischer) for 2 hours at 37°C in a water bath under mild shaking. Control samples were incubated with the same amount of bioparticles and for the same time at 4°C to inhibit phagocytosis. All samples were then subjected to RBC lysis. After washing, cells were acquired on a FACS Lyrics cytometer. Results are expressed as the ratio of pHRodo mean fluorescent intensity (MFI) from 37°C-incubated samples to 4°C-incubated control samples.

### Oxidative burst assay

Diluted heparinized whole blood (1/10) was pre-incubated with 600ng/ml dihydroethidium (DHE, Sigma-Aldrich) probe for 15 min at 37°C under agitation in a water bath before being primed with TNFα (5ng/ml, Bio-techne), LPS (10ng/ml, Sigma-Aldrich), or TLR7/8 agonist CL097 (2.5µg/ml, Invivogen) for 45 min at 37°c. Samples were then stimulated with N-formyl-methionyl-leucyl-phenylalanine (fMLP 1µM, Sigma-Aldrich) for 5 min and subjected to RBC lysis (BD BioSciences) and washing before acquisition on a FACS Lyrics cytometer. Results were expressed in MFI and a stimulation index (SI) was calculated (MFI ratio between stimulated and unstimulated cells).

### NETosis assay

Neutrophils were purified from EDTA-treated whole blood using MACSXpress Neutrophil isolation kit (Miltenyi Biotech) which allowed a purity routinely over 98%. Purified neutrophils were suspended at 1×10^6^ cells/mL in Sytox Green solution (Thermofischer, final concentration 2.5μM) diluted in Hank’ s Balanced Salt (HBSS) and preincubated for 30 min with either 5 ng/ml TNF-α, 10 ng/ml lipopolysaccharide (LPS, Sigma), 2.5µg/ml CL097 (Invivogen), or medium. Then, 150μL of cells were seeded in black 96-well plates and stimulated with either 25nM phorbol myristate acetate (PMA, Sigma), 1µM fMLP, or 5μg/ml *S*.*aureus* peptidoglycan (PGN, Sigma-Aldrich) for 3h at 37°C. Cell-free DNA release was quantified over time on an Infinite 200 PRO microplate reader (TECAN). Results were expressed as the increase in fluorescence over 3h normalized for baseline.

### Quantification of circulating activation markers

Plasma and serum were obtained by centrifugation of fresh blood and immediately frozen at - 80°C after aliquoting.

Soluble CD14 (sCD14), lactoferrin, L-selectin, matrix metallopeptidase 9 (MMP-9), neutrophil gelatinase-associated lipocalin (NGAL), IL-8 (CXCL8), myeloperoxidase (MPO) and S100A8/A9 (calprotectin) were quantified in the plasma using Procartaplex Multiplex Immunoassay (Thermofisher Scientific) according to the manufacturer’ s instructions. Plates were read on a MAGPIX System (Merck Millipore) analyzer and data analyzed with Milliplex software. Human neutrophil elastase plasma levels were quantified by ELISA using Human PMN-Elastase ELISA kit (ThermoFisher Scientific) according to manufacturer’ s instructions. Absorbance was read at 450nm using a Multiskan FC spectrophotometer (ThermoFischer).

Neutrophil Extracellular Traps (NETs) were quantified by measuring myeloperoxidase (MPO)-DNA complexes in serum samples using an in-house capture ELISA already described *(61, 62)*. Samples were interpolated from a standard curve and results were expressed in arbitrary units (a.u.). The detection range was 4.2-724 a.u. Values below the lower limit of detection (LDD) were assigned the LDD/2 (i.e. 2.1 a.u.) and values above the maximal of standard curve (MSC) were assigned the 1.5 X MSC (i.e. 1086 a.u).

### Statistics

Intergroup differences were analyzed with non-parametric unpaired Mann-Whitney U-test for comparison between two groups and Kruskall-Wallis test followed by Dunn post-test for comparison between more than two groups. Correlation between continuous data was performed using Spearman correlation. Hierarchical clustering was performed using Euclidian distance and Ward linkage method. Statistical tests were bilateral, and a type I error was fixed at 5%. Statistical analyses were performed with GraphPad Prism versions 8.0 (GraphPad Software Inc.), Statview 5.0 (SAS Institute Inc.) and hierarchical clustering with Genesis 1.8.1 (Gratz University of Technology).

## Data Availability

All data is available upon reasonable request

## Supplementary Materials

Fig. S1. **Plasma concentration of neutrophil elastase and matrix metalloproteinase-9**

Fig. S2. **Neutrophil subpopulations analysis**.

Fig. S3. **Monocyte subpopulations analysis**

Fig. S4. **Monocyte subpopulation and mortality**

Fig. S5. **Soluble markers and mortality**

Table S1. **Antibody list**.

## Acknowledgments

We thank Nicolas Heddebaut for technical assistance with multiplex testing and Stephane Ruckly for data extraction.

## Funding

This work was funded by the French National Research Agency (ANR) RA-COVID-19 V5 grant under the name COVINNATE.

## Author contributions

Conceptualization: LdC, MHN, SCM, RM

Investigation: MP,VG, LDC, MHN, PHW, DKT, JFT, FXL, YY, ATD, PM

Visualization: MP, LdC, VG

Funding acquisition: MHN, LdC

Supervision: LdC, SCM, MHN

Writing – original draft: LdC, MP, VG, SCM

Writing – review & editing: All authors

## Competing interests

Authors declare that they have no competing interests.

## Data and materials availability

All data are available in the main text or the supplementary materials.

## Notes

### Competing Interest Statement

The authors have declared no competing interest.

### Author Declarations

The study was approved by National Ethics committee CEEI/IRB under the number 20-715.

