## Supplementary Material for "Innate immune deficiencies in patients with COVID-19"

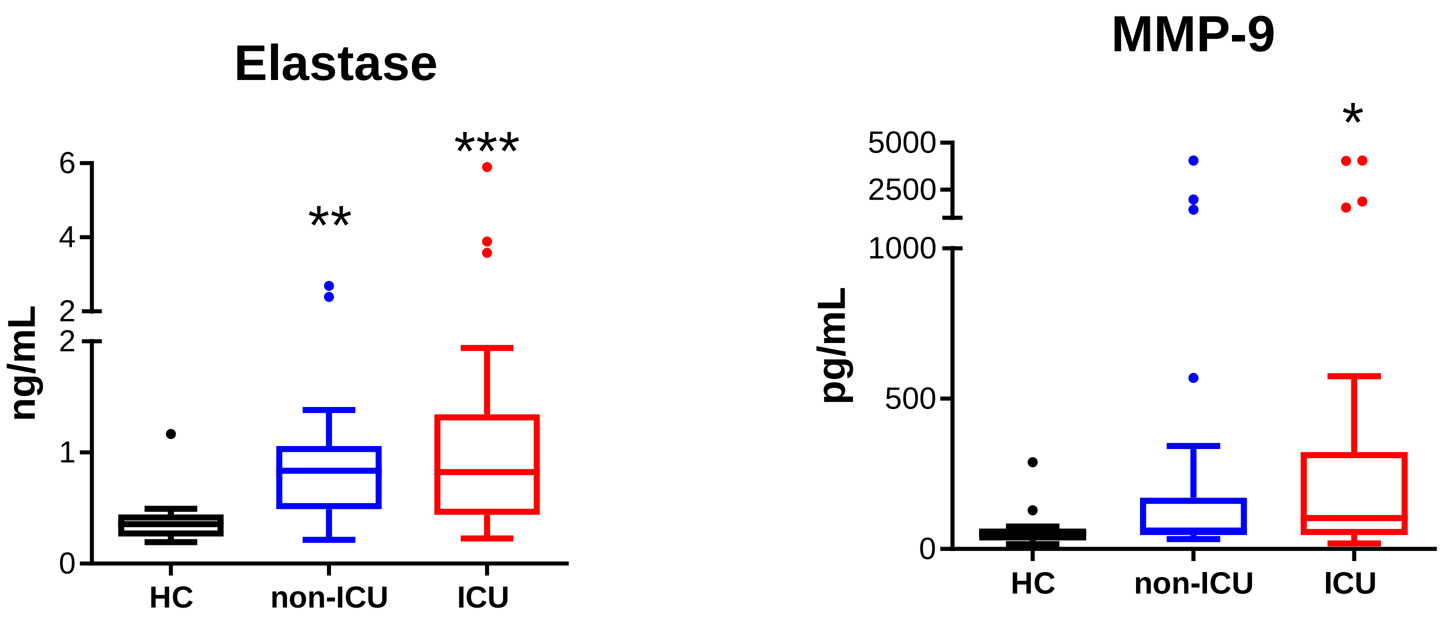


**Supplementary Figure 1**

**Plasma concentration of neutrophil elastase and matrix metalloproteinase-9.**
Intergroup comparison by Mann-Whitney U test between healthy controls and ICU or non-ICU patients: ***P < 0.001, **P < 0.01, *P < 0.05 All boxplots whiskers represent 10$^{th}$ and 90$^{th}$ percentiles. HC: healthy controls

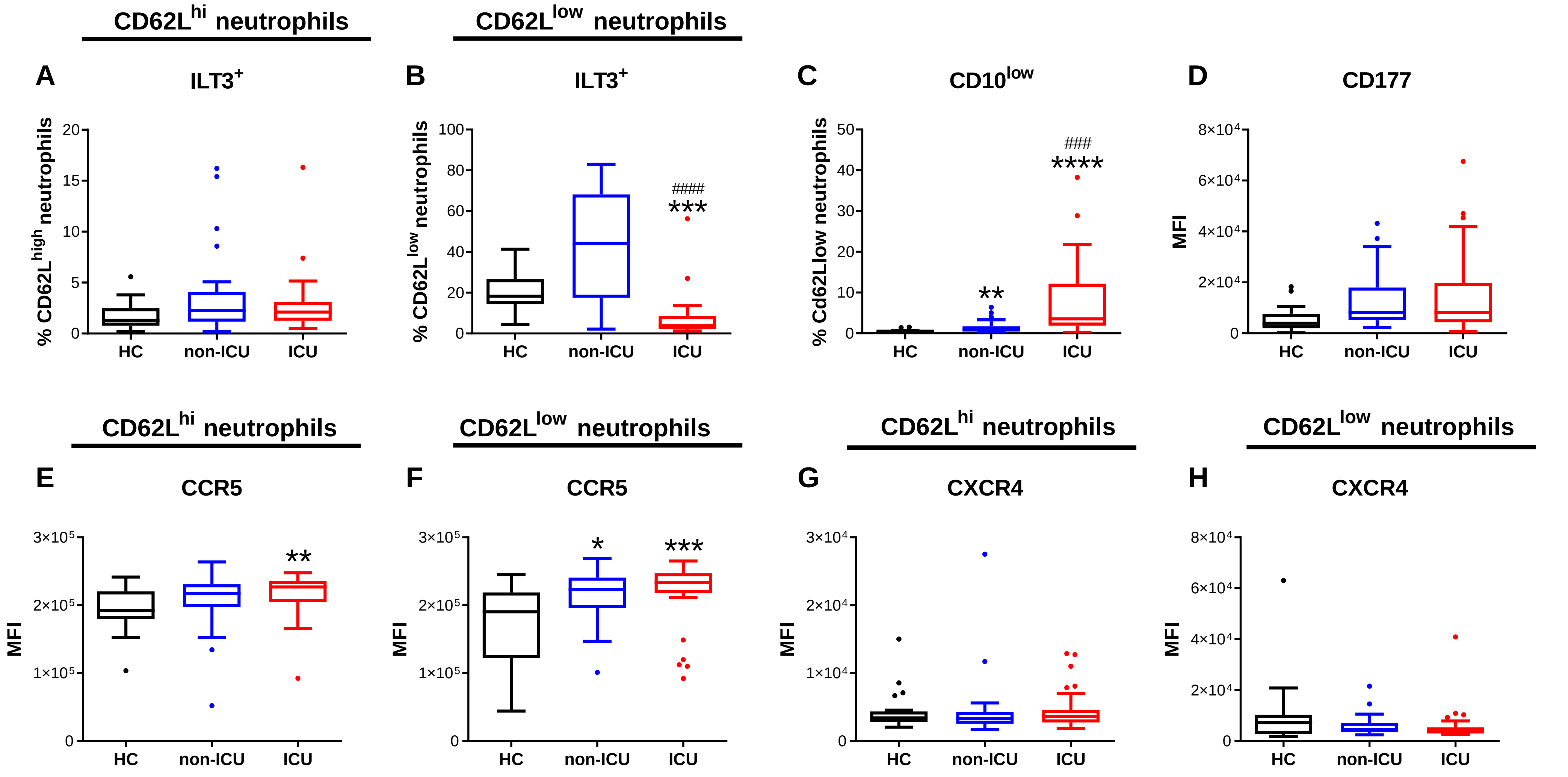


**<u>Supplementary Figure 2</u>. Neutrophil subpopulations analysis**

**(A-B)** Percentage of ILT3+ cells in (A) CD62Lhi neutrophils and (B) CD62Llow neutrophils. **(C)** Percentage of CD10low cells on CD62Llow neutrophils. **(D)** Expression of CD177 on total neutrophils. **(E-F)** Expression of CCR5 on (E) CD62Lhi and (F) CD62Llow neutrophils. **(G-H)** Expression of CXCR4 on (G) CD62Lhi and (H) CD62Llow neutrophils. Intergroup comparison by Mann-Whitney U test between healthy controls and ICU or non-ICU patients: ****P < 0.0001, ***P < 0.001, **P < 0.01, between non-ICU and ICU patients: ###P < 0.001. All boxplots whiskers represent 10th and 90th percentiles. HC: healthy controls.

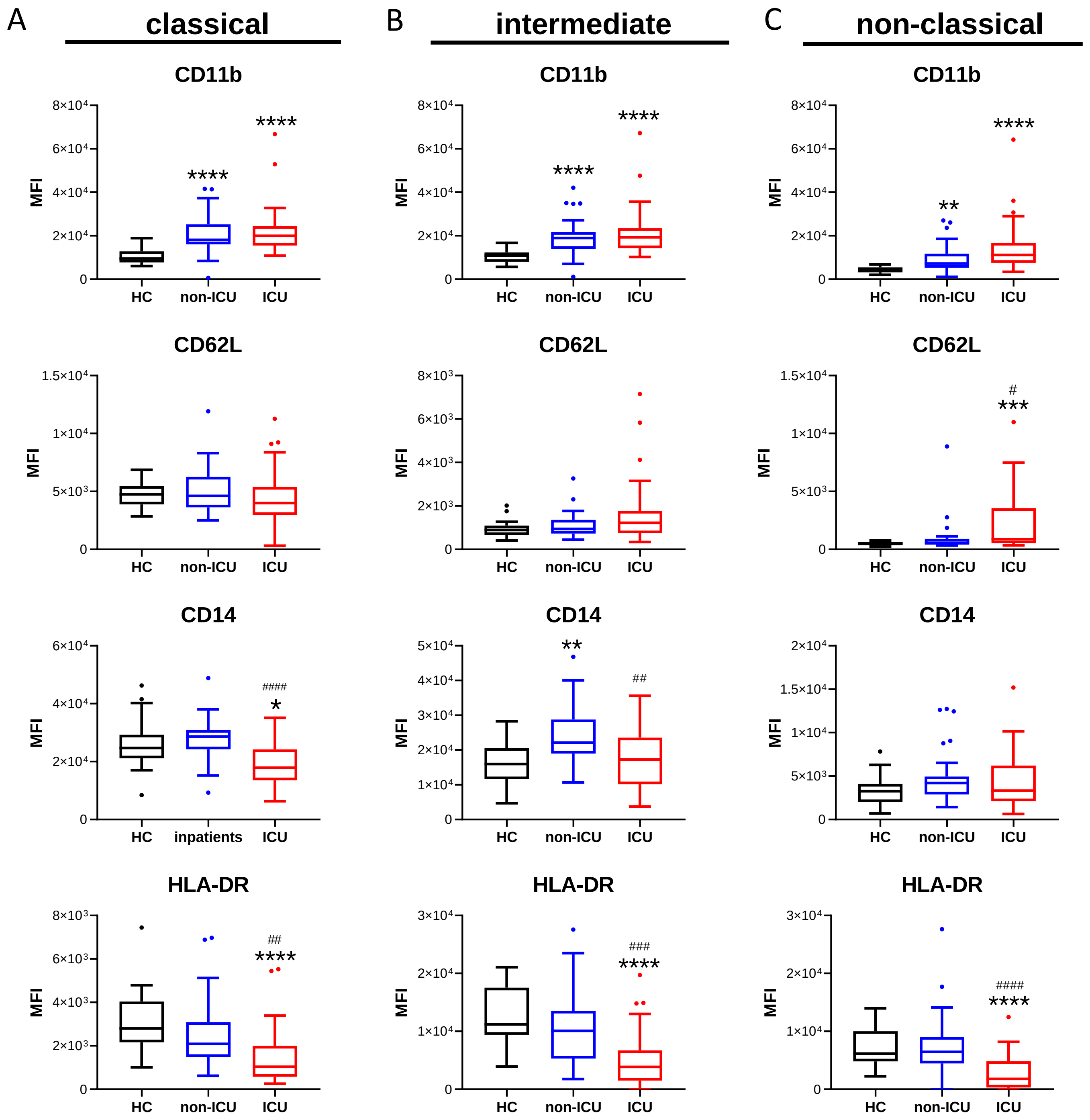


**Supplementary Figure 3.**

**Monocyte subpopulations analysis**

**(A-C)** Expression of activation markers CD11b, CD62L, CD14 and HLA-DR on (A) CD14+CD16- classical monocytes, (B) CD14+CD16+ intermediate monocytes, (C) -CD16+ atypical monocytes. Intergroup comparison by Mann-Whitney U test between healthy controls and ICU or non-ICU patients: ****P < 0.0001, ***P < 0.001, **P < 0.01, *P < 0.05, between non-ICU and ICU patients: ####P < 0.0001, ###P < 0.001, ##P < 0.01, #P<.05.. All boxplots whiskers represent 10th and 90th percentiles. HC: healthy controls.

### Supplementary Figure 4

**Monocytes subpopulation and mortality** Phenotypic markers significantly different between patient who survived (red) or died (black) during their ICU stay on classical monocytes (top row), intermediate monocytes (center row), atypical monocytes (bottom row, left) and non-classical (CD16+) monocytes. The histograms represent the median and the error bars the interquartile range. Intergroup comparison by Mann-Whitney U test between survivors and deceased patients, **P < 0.01, *P < 0.05

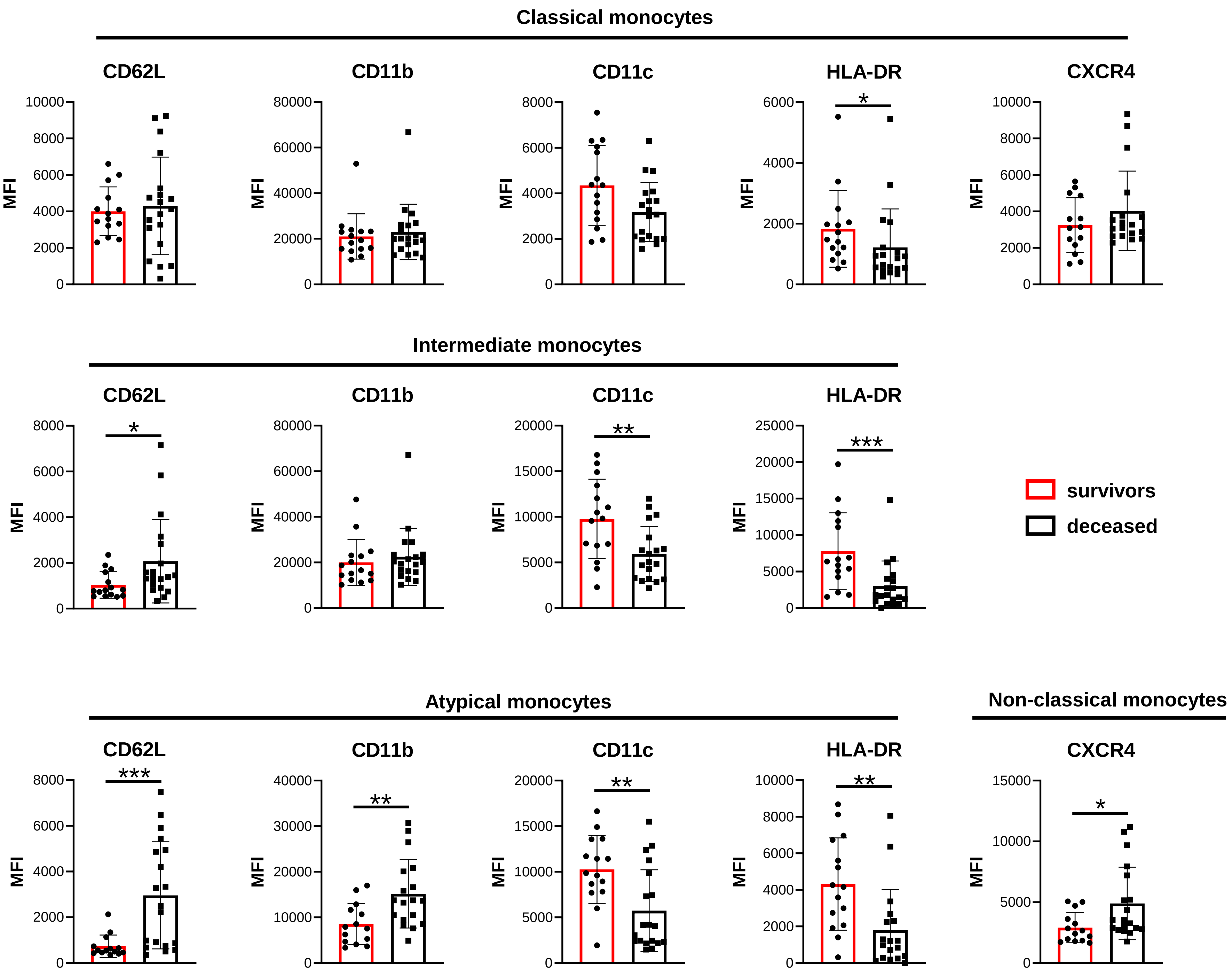

**Supplementary Figure 5**

**Soluble markers and mortality**

Soluble markers significantly different between patient who survived (red) or died (black) during their ICU stay in plasma. The histograms represent the median and the error bars the interquartile range. Intergroup comparison by Mann-Whitney

*P < 0.05

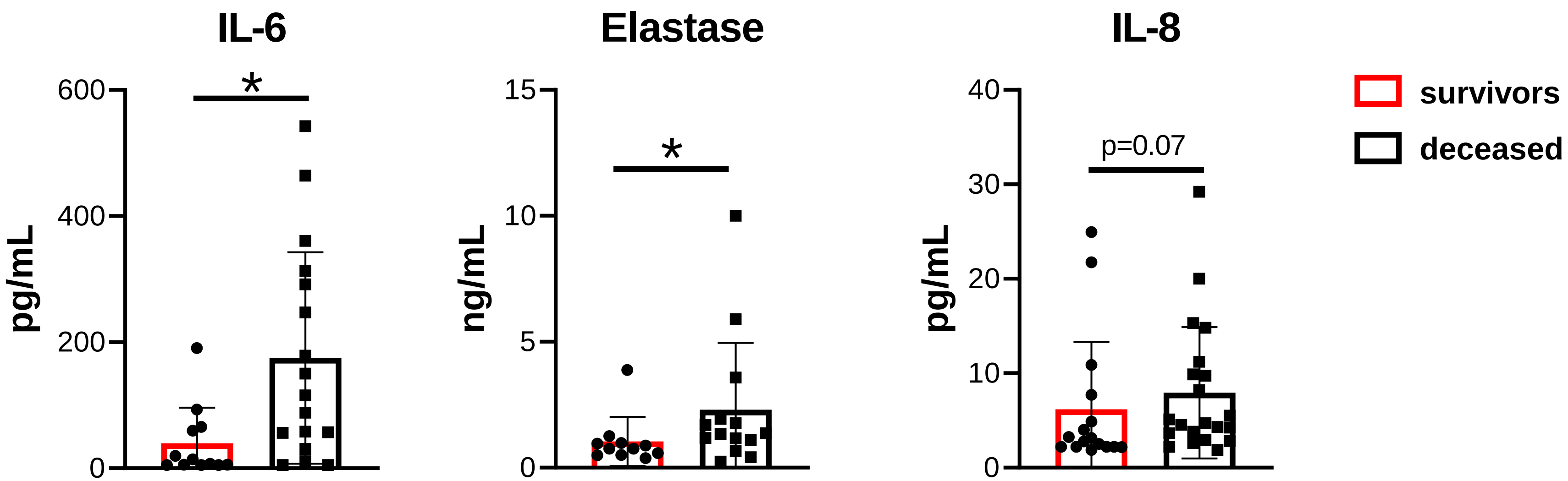

Table S1. **Antibody list**.

| Target | Clone | Provider |
|---|---|---|
| CD66b | G10F5 | Biolegend |
| CD11b | ICRF-44 | BD Biosciences |
| CD62L | DREG-56 | BD Biosciences |
| CD16 | 3G8 | Biolegend |
| CD14 | MϕP9 | BD Biosciences |
| HLA-DR | G46-6 | BD Biosciences |
| LOX-1 | 15C4 | Biolegend |
| PD-L1 | MIH3 | Biolegend |
| CD10 | HI10a | Biolegend |
| CD13 | WM15 | Biolegend |
| ILT3 | ZM4.1 | Biolegend |
| CD49d | 9F10 | Biolegend |
| CD195 | J418F1 | Biolegend |
| CD184 | 12G5 | Biolegend |
| CD33 | WM53 | Biolegend |
| TLR2 | 11G7 | BD Biosciences |
| TLR4 | TF901 | BD Biosciences |
| TLR7 | 9L859 | MyBioSource |
| TLR8 | S16018A | Biolegend |
| Dectin-1 | 15E2 | Biolegend |
| RIG-1 | 4G1B6 | ThermoFisher |
| MDA-5 | 33H12L34 | ThermoFisher |
| PKR | YE350 | Abcam |
| MAVS | ABM28H9 | ThermoFisher |
| pNFkB p65 (Ser529) | B33B4WP | eBioscience |
| pIRF3 (Ser386) | E7J8G | Cell Signaling Technology |
| pIRF7 (S477-S479) | K47-671 | BD Biosciences |
| CD177 | MEM-166 | Biolegend |
| CD11c | S-HCL-3 | Biolegend |
| CCR2 | K036C2 | Biolegend |
| CD36 | 5-271 | Biolegend |
| TCRαβ | IP26 | Biolegend |
